# Differential Clearance of [^177^Lu]Lu-PSMA-617 from Metastatic Prostate Cancer Lesions and Normal Tissues creates a Window for Safe Application of DNA Repair Inhibitors

**DOI:** 10.1101/2025.10.08.25337591

**Authors:** James Russell, Milan Grkovski, Joseph A. O’Donoghue, Alexandre Franca-Velo, Simone Krebs, Lisa Bodei, Heiko Schöder, John L. Humm

## Abstract

**Background:** DNA repair inhibitors may safely enhance targeted radiotherapy if they can be administered when radioisotope has cleared from critical normal tissues. We investigated whether the kinetics of [^177^Lu]Lu-PSMA-617 (Pluvicto) would be consistent with such a strategy in metastatic castrate resistant prostate cancer (mCRPC).

**Materials and Methods:** Pluvicto effective t_1/2_ was measured for organs-at-risk (kidneys, parotid and submandibular glands) and lesions from 60 mCRPC patients. Based on this, a safe strategy for DDR inhibition was defined, and its potential benefit to the lesion was calculated, using published *in vitro* radiosensitivity data.

**Results:** Pluvicto clearance is almost 3× faster in normal organs-at-risk compared to lesions. Using pre-clinical estimates of DDRi sensitization, a meaningful increase in lesion effective treatment dose could be achieved with minor additional risk to normal organs.

## INTRODUCTION

[^177^Lu]-LuPSMA-617 (Pluvicto) has enhanced progression-free survival in metastatic castration resistant prostate cancer (mCRPC), with only minor benefits for long term survival (1). It might be possible to improve outcome with the use of DNA repair inhibitors (DDRi), which produce dramatic radio-sensitization in vitro and in vivo. However, their effects are not confined to tumor tissue (2), and thus their application, combined with external beam radiotherapy creates potentially serious risks of unacceptable normal tissue damage (3). For Pluvicto or any targeted radiotherapy, there is a conceptually simple way to apply a DDRi: when a radiopharmaceutical is cleared from the dose-limiting normal tissues faster than from the target lesions, a safe time is created, during which administration of a radiosensitizer poses no risk.

It is of course an oversimplification to think of a radiopharmaceutical falling to zero in normal tissue, as there will always be some retained activity. Applying the radiosensitizer when (for example) the dose limiting normal tissue has absorbed 99% of its total dose implies increasing the damage from the residual activity. The level of toxicity will depend on the degree of sensitization that the repair inhibitor actually produces in clinical practice, and a slight increase in normal tissue risk is unavoidable.

In this report, we present data on the clearance of Pluvicto from mCRPC as well as from the dose-limiting tissues (kidneys and salivary glands) and find that a time window exists when repair inhibitors can be given with minimal impact on normal tissues. We then apply the existing *in vitro* data on DDRi repair inhibition to discuss the range of potential benefits that might be obtained

## METHODS

Sixty mCRPC patients underwent triple time-point SPECT/CT imaging after receiving 7.3±0.3 GBq of Pluvicto. Imaging acquisition and dosimetry were described previously (4). For N=8 patients, the salivary glands were not within the field of view. Effective t_1/2_ for both N=1200 lesions and normal organs was calculated via mono-exponential fitting of the decay-corrected time-activity curves.

## RESULTS AND DISCUSSION

A representative SPECT/CT image set is shown in Figure 1. Compared to the lesions, activity is rapidly lost from the kidneys. This pattern was consistent throughout the patient group (Table 1). The distribution of effective half-lives is shown in Figure 2A. Figure 2B shows the inter-patient variation for lesion effective half-life.

**TABLE 1.** Effective half-life of Pluvicto in lesions and normal tissues

| Tissue | N | Median (h) | Interquartile range (h) | Coefficient of variation (%) |
| --- | --- | --- | --- | --- |
| Lesion | 1200 | 86.7 | 59.3 | 40 |
| Kidney | 60 | 29.8 | 8.6 | 24 |
| Submandibular gland | 52 | 31.8 | 9.9 | 26 |
| Parotid gland | 52 | 32.1 | 10.7 | 25 |

**Figure 1.**
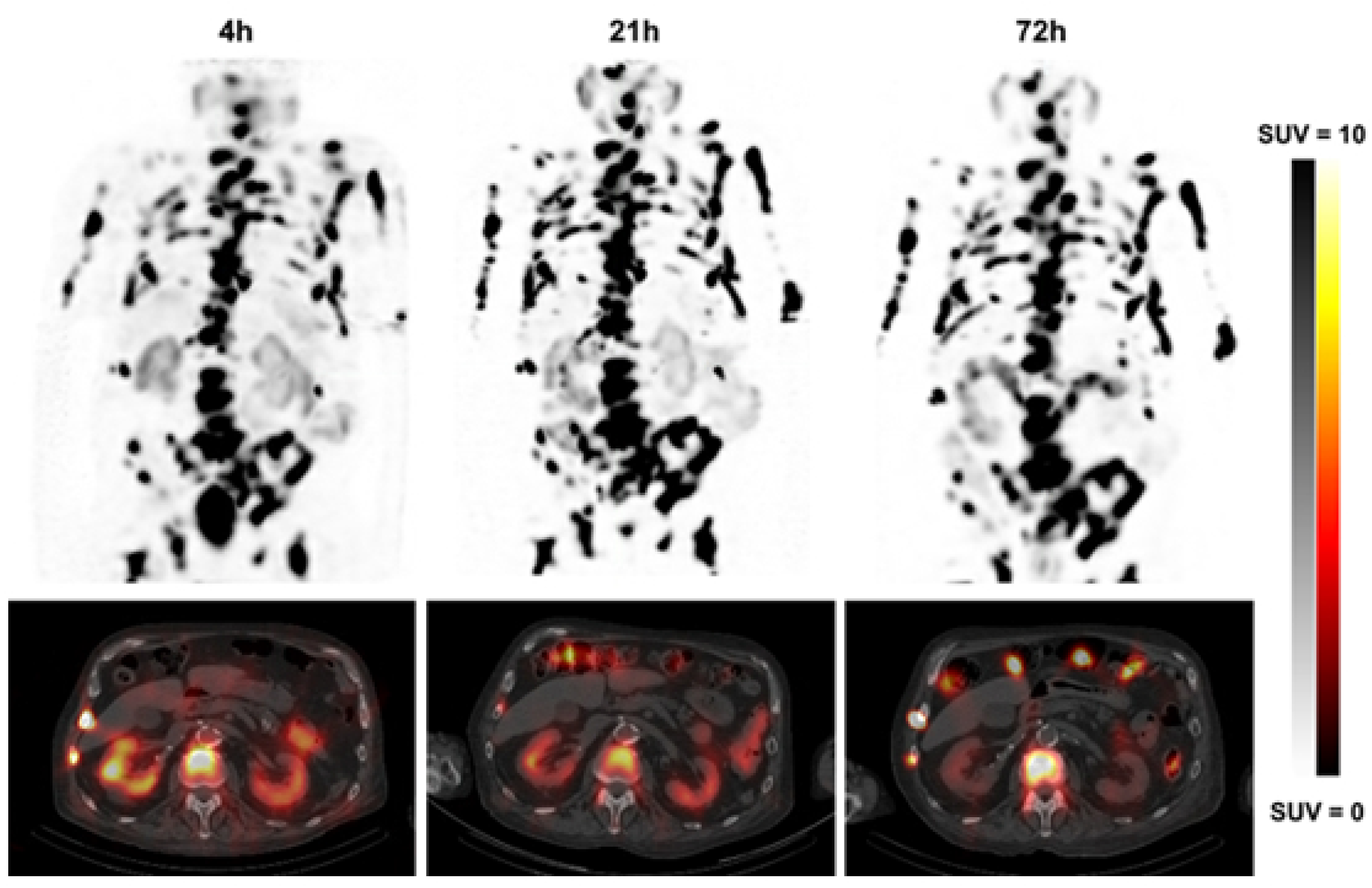
Serial SPECT/CT of an 82 yr old mCRPC patient with baseline PSA = 607 ng/ml. Effective t_1/2_ were 26.6 h (kidneys) and 156 h (median from N=40 lesions).

**Figure 2.**
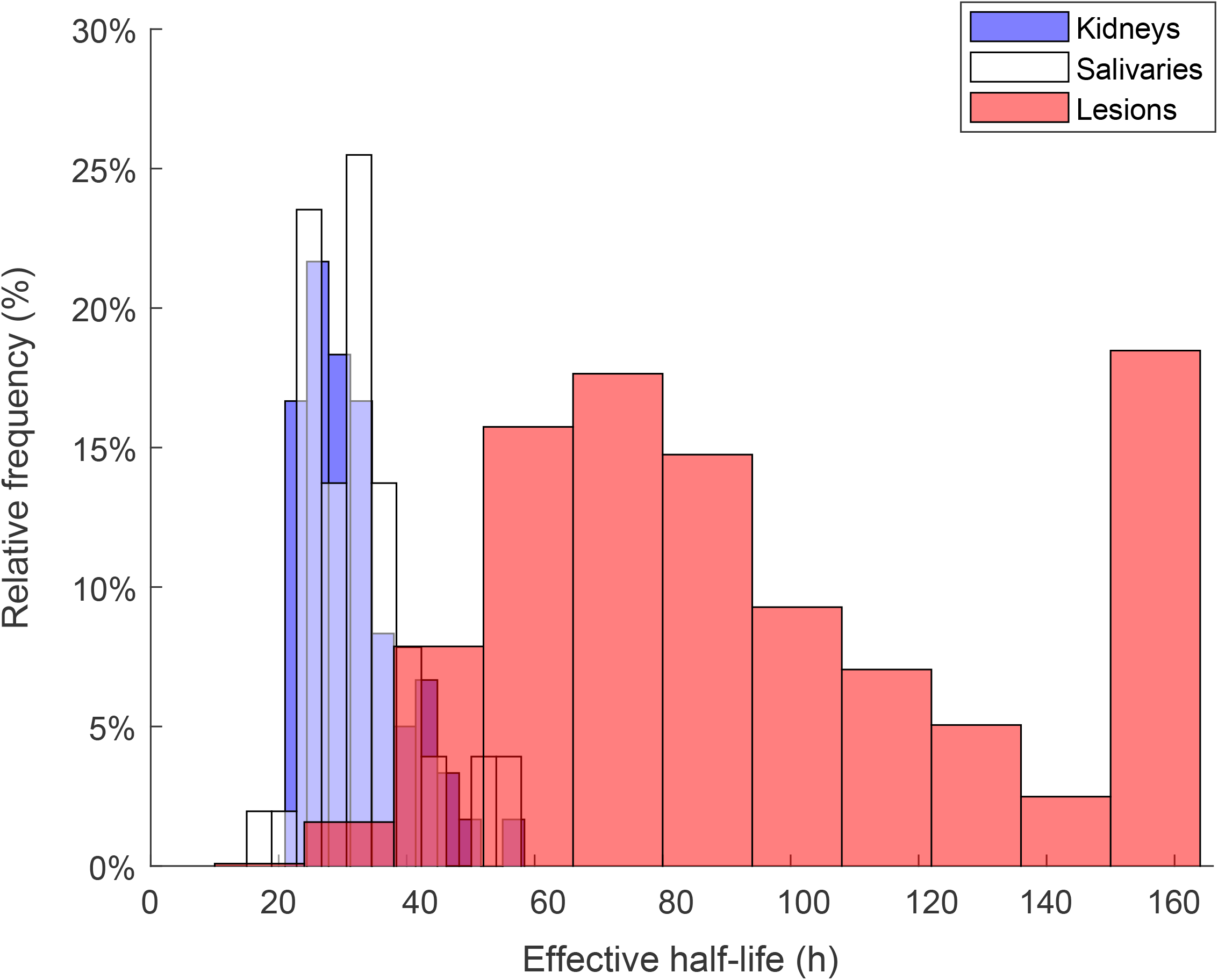
**A)** Distribution of Pluvicto effective half-life values in lesions and normal tissues. **B)** The median effective half-life and inter-quartile range for patients with **≥**5 lesions.

Figure 3A demonstrates that, across lesions, Pluvicto retention shows only a weak correlation with absorbed dose. Instead, the major determinant of lesion dose is the absolute uptake of Pluvicto, as displayed in Figure 3B. This is important, since the lesions that will benefit most from our proposed strategy are those with the most protracted retention of activity. If there were a correlation between Pluvicto half-life and lesion dose, the lesions receiving the least dose would also receive the least sensitization. Using total lesion volumetric intensity (Gy*cc) as a surrogate for overall disease burden, we can see that prolonged Pluvicto retention is not confined to individuals with low tumor burden (Figure 4).

**Figure 3.**
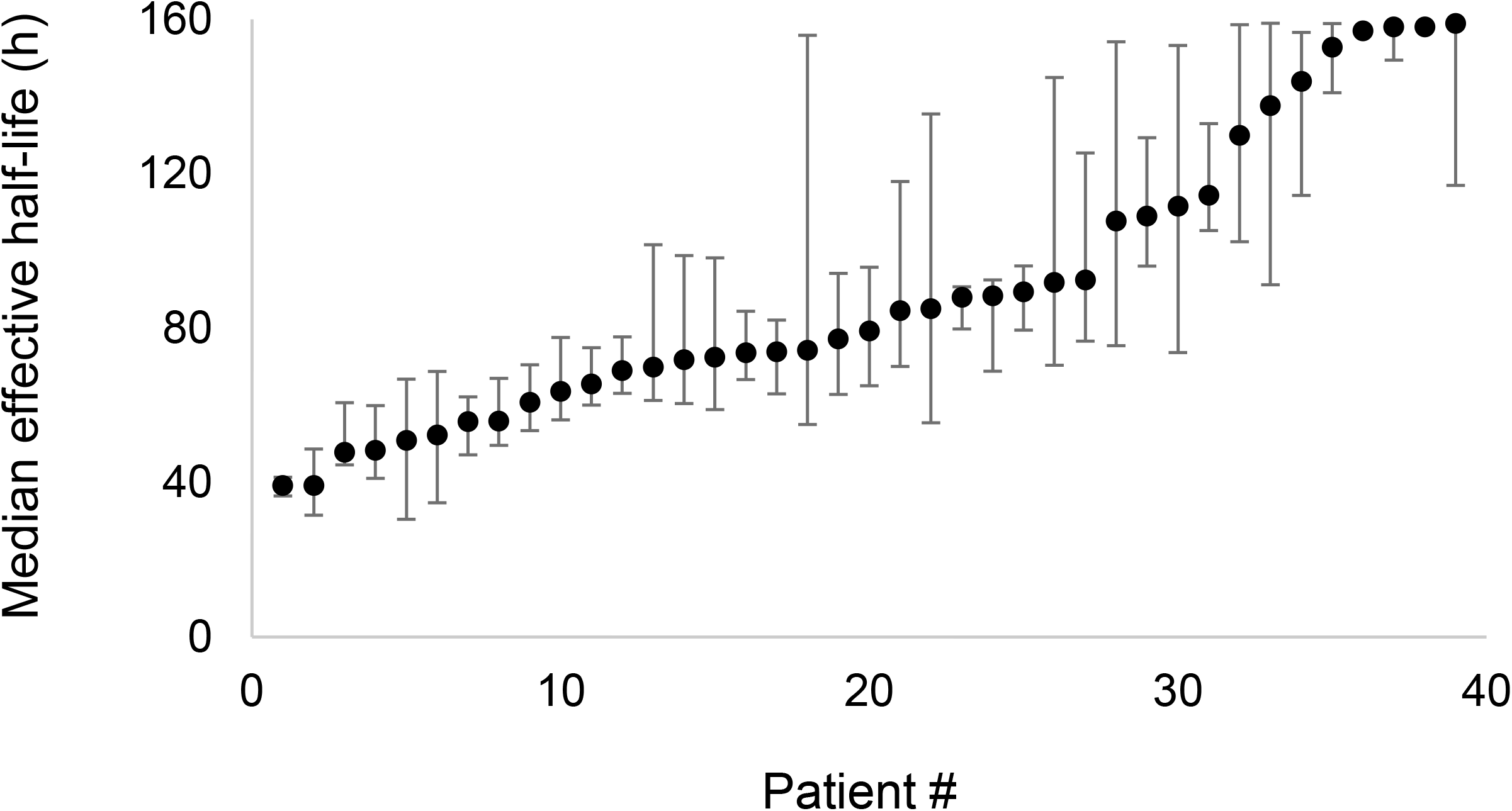
Relationship between **A)** effective half-life of Pluvicto and the lesion dose for 1200 lesions and **B)** SUV_max_ 24 hours post injection and lesion dose.

**Figure 4.**
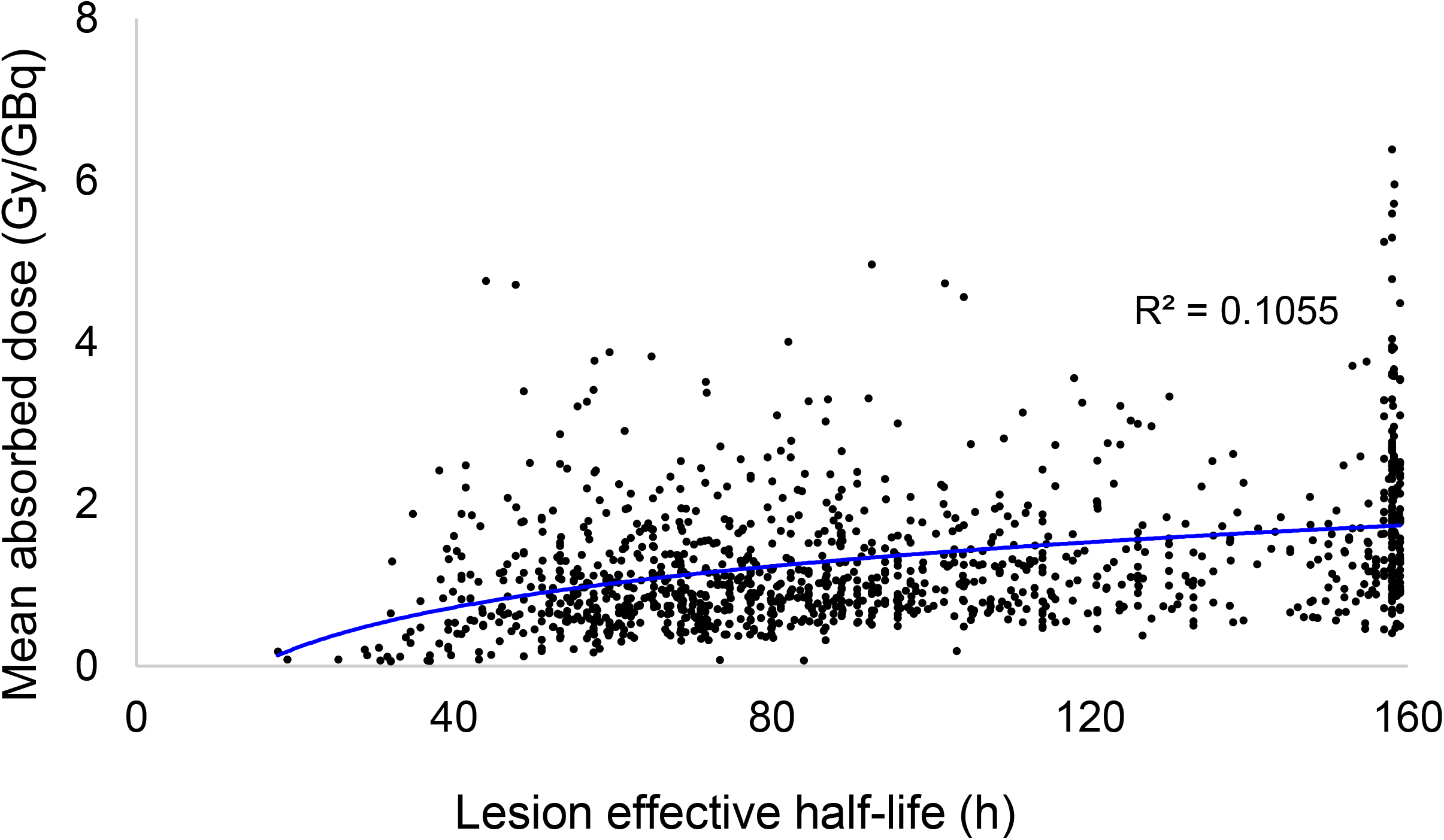
The relationship between total lesion volumetric intensity product and median effective half-life on a per-patient basis, by logarithmic regression.

To calculate the risks and benefits of administering a DDRi, we use the published data on the ATMi inhibitor AZD1390 and survival curves of fibroblasts from Ataxia-Telangiectasia (AT) individuals. The magnitude of radio-sensitization is given by the sensitization enhancement ratio (SER), defined as thethe ratio of radiation doses required for some specified effect in the presence and absence of the sensitizer. The literature suggests that, for AZD1390, a plausible range for the SER could be between 2 – 5 (5-7). These studies were conducted at dose rates typical of external beam radiotherapy; regarding lower dose rates relevant to radiopharmaceutical therapy, the only available data comes from experiments with AT fibroblasts, which indicates a very similar range (8-11).

At 195h after Pluvicto administration, ∼1.0% and 1.6% of the initial activity would remain in the kidneys and parotid glands, respectively. Applying a sensitizer at this point would, even under the most extreme assumption (five-fold sensitization), this would correspond to a modest increase, equivalent to ∼6.4% more activity in the parotids.

For comparison, 21% of the initial activity would remain in a lesion with a Pluvicto effective t_1/2_ of 86.7 h. Assuming that DDR inhibition could produce sensitization between 2 and 5-fold (in terms of dose required for iso-effect), the median lesion would receive a boost equivalent to a 23–84% increase in the initial activity. Even greater sensitization would occur in lesions with longer radiopharmaceutical retention: Figure 5 shows how the expected benefit varies with the half-life of Pluvicto in the lesion and with the degree of sensitization. Thus, the upper and lower lines represent the bounds of what could reasonably be expected to occur. A further opportunity can be deduced from the results of Karimzadeh et al (12): lesions – though not normal tissue -receive their greatest dose during the first Pluvicto cycle. For a given target of normal tissue risk, confining sensitization to cycle 1 would further enhance the tumor treatment. Finally, the normal tissue absorbed dose from Pluvicto can be substantially reduced by proper hydration (4), and this may allow earlier administration of a DDRi, and thus greater sensitization.

**Figure 5.**
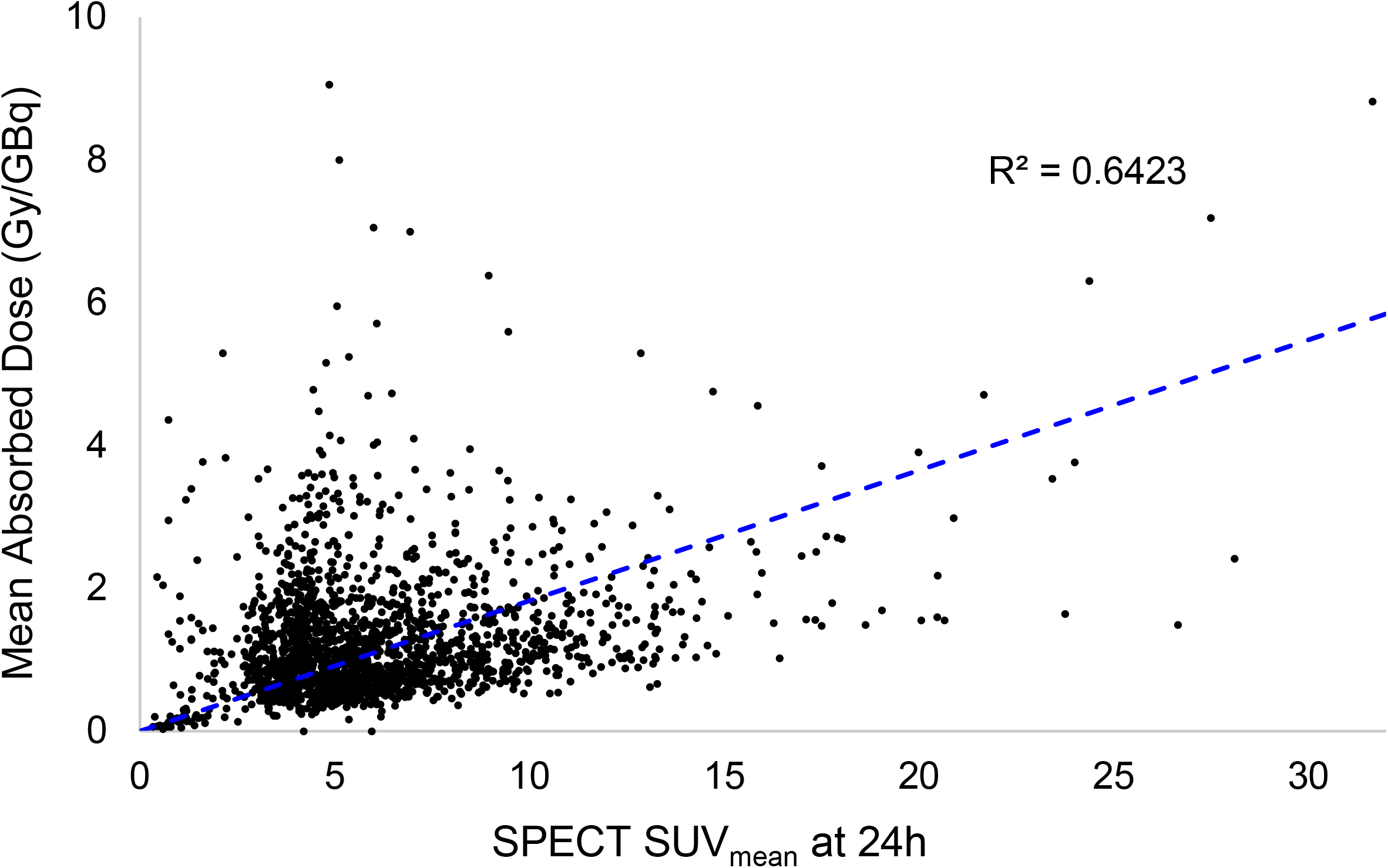
The potential benefit applying a DDRi at 195 hours after Pluvicto administration. Benefit was calculated by multiplying the residual dose at 195 hrs by the SER, and adding to the dose already delivered.

The main concern with DDRi in this context is the requirement for continuous inhibition over multiple days. This may be possible: for example, AZD1390 achieves a maximum human plasma concentration of 20 nM (13), and has a plasma half-life of ∼10 hours (14), but is radio-sensitizing at 10 nM (15). In addition to the ATM protein, another potential target is the DNA dependent protein kinase, and there are multiple inhibitors to be compared for the greatest utility (16).

In conclusion, our analysis suggests it is possible to envisage a reasonably safe way of incorporating DDR inhibitors into radiopharmaceutical therapy. Such a strategy could benefit most patients, with a smaller subset being particularly helped.

## Data Availability

All data produced in the present study are available upon reasonable request to the authors

---

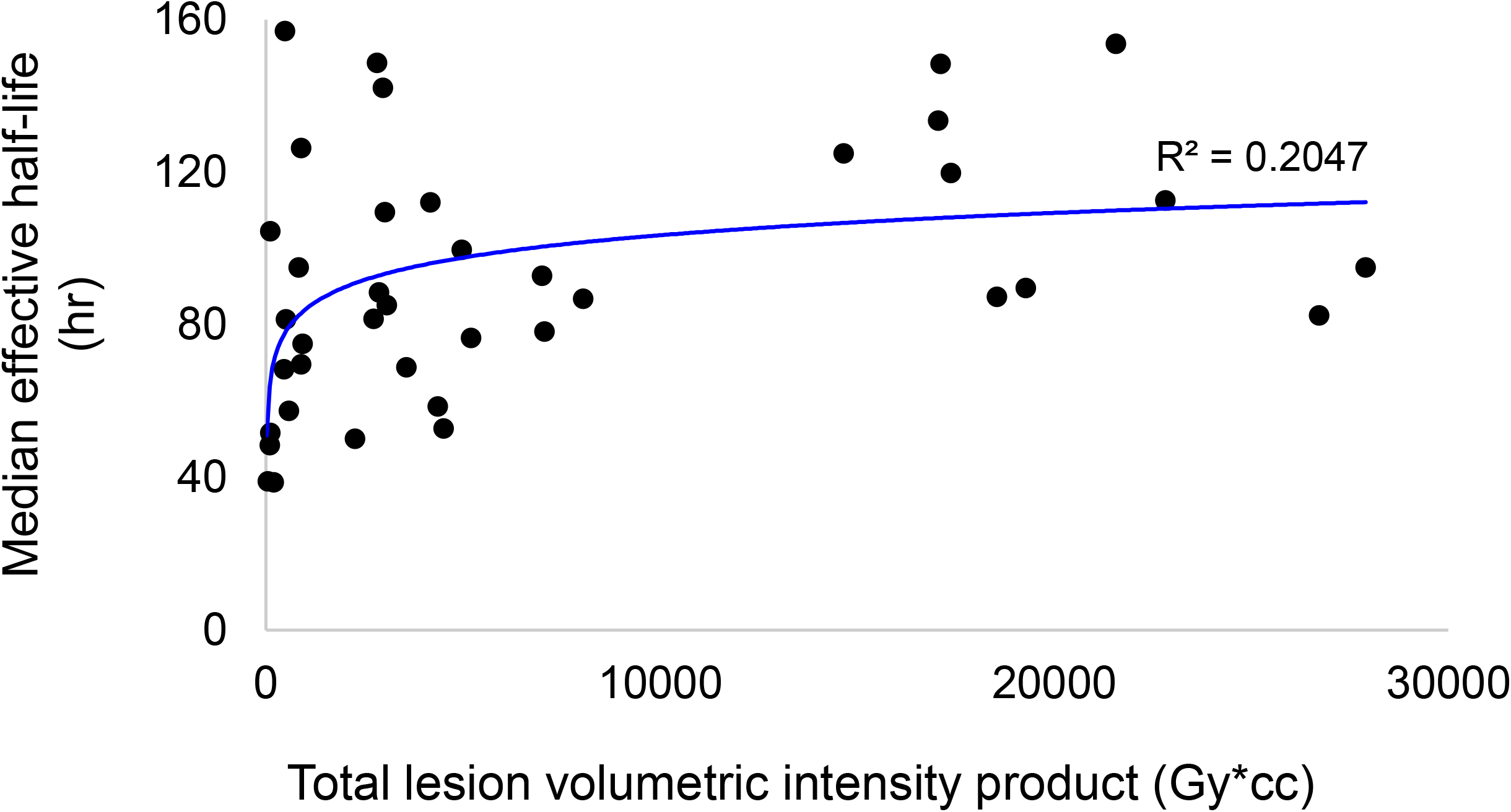

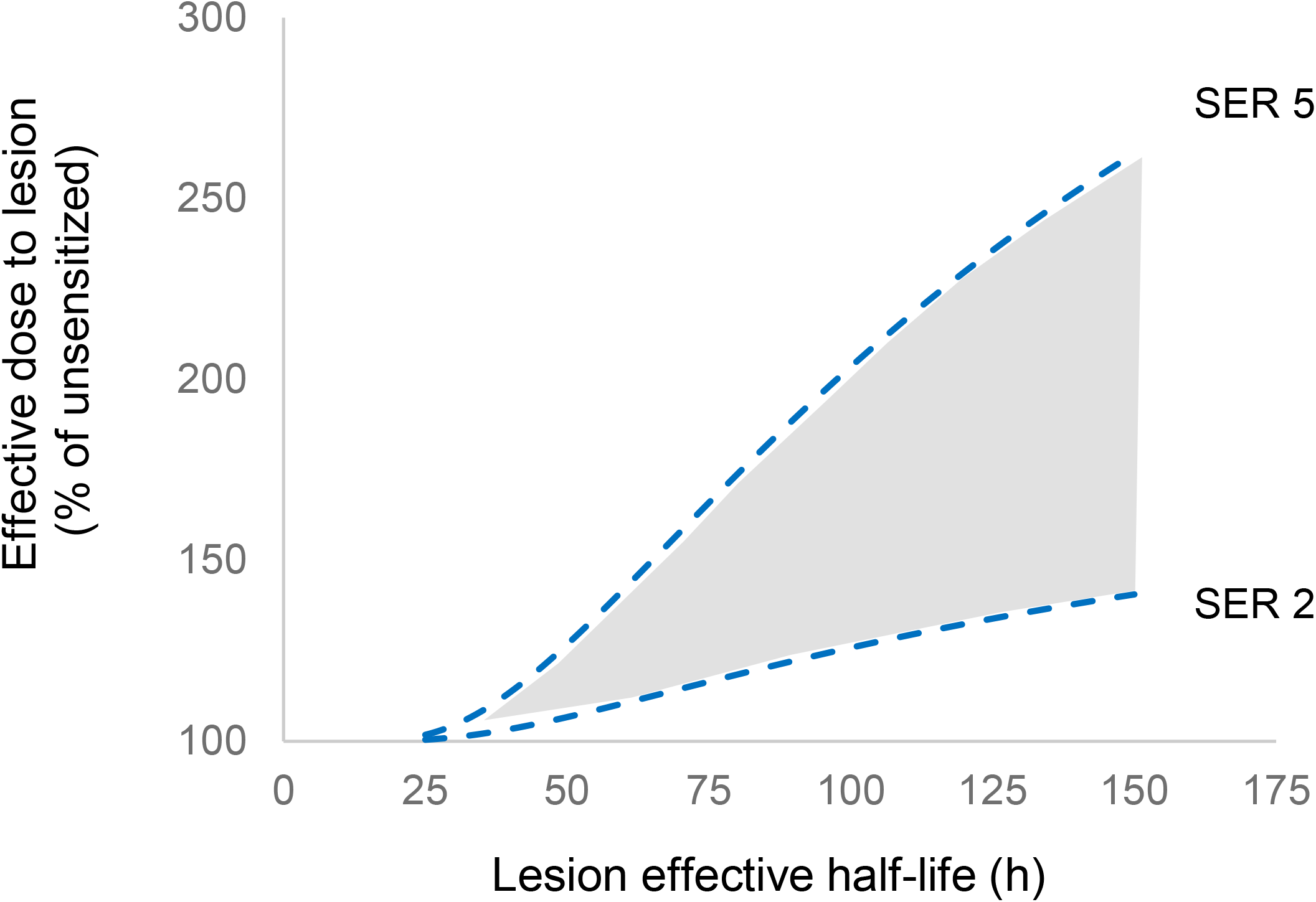

## Notes

Research at Memorial Sloan Kettering Cancer Center is supported by NIH/NCI Cancer Center support grant P30 CA008748.

### Competing Interest Statement

Research at Memorial Sloan Kettering Cancer Center is supported by NIH/NCI Cancer Center support grant P30 CA008748. Simone Krebs is supported in part by NIH grant R37 CA262557 and has consulted for Telix Pharmaceuticals Ltd. Lisa Bodei reports nonremunerated consultancies from Novartis, ITM, Ipsen, Iba, Great Point Partners, PointBiopharma, RayzeBio, Abdera, Fusion, SolveTx, and Wren Laboratories and an institutional grant from Novartis. No other potential conflicts of interest relevant to this article exist.

### Funding Statement

Research at Memorial Sloan Kettering Cancer Center is supported by NIH/NCI Cancer Center support grant P30 CA008748. Simone Krebs is supported in part by NIH grant R37 CA262557.

### Author Declarations

This study was conducted following the principles outlined in the Declaration of Helsinki and approved by Memorial Sloan Kettering Cancer Center's Institutional Ethics Committee, which waived the requirement for study-specific consent.

